# Across-Frequency Temporal Processing and Speech Perception in Cochlear Implant Recipients and Normal Hearing Listeners

**DOI:** 10.1101/2020.07.21.20159160

**Authors:** Chelsea M. Blankenship, Jareen Meinzen-Derr, Fawen Zhang

## Abstract

**Objective:** Individual differences in temporal processing contributes strongly to the large variability in speech recognition performance observed among cochlear implant (CI) recipients. Temporal processing is traditionally measured using a behavioral gap detection task, and therefore, it can be challenging or infeasible to obtain reliable responses from young children and individuals with disabilities. Within-frequency gap detection (pre- and post-gap markers are identical in frequency) is more common, yet across-frequency gap detection (pre- and post-gap markers are spectrally distinct), is thought to be more important for speech perception because the phonemes that proceed and follow the rapid temporal cues are rarely identical in frequency. However, limited studies have examined across-frequency temporal processing in CI recipients. None of which have included across-frequency cortical auditory evoked potentials (CAEP), nor was the correlation between across-frequency gap detection and speech perception examined. The purpose of the study is to evaluate behavioral and electrophysiological measures of across-frequency temporal processing and speech recognition in normal hearing (NH) and CI recipients.

**Design:** Eleven post-lingually deafened adult CI recipients (n = 15 ears, mean age = 50.4 yrs.) and eleven age- and gender-matched NH individuals participated (n = 15 ears; mean age = 49.0 yrs.). Speech perception was evaluated using the Minimum Speech Test Battery for Adult Cochlear Implant Users (CNC, AzBio, BKB-SIN). Across-frequency behavioral gap detection thresholds (GDT; 2 kHz to 1 kHz post-gap tone) were measured using an adaptive, two-alternative, forced-choice paradigm. Across-frequency CAEPs were measured using four gap duration conditions; supra-threshold (behavioral GDT x 3), threshold (behavioral GDT), sub-threshold (behavioral GDT/3), and reference (no gap) condition. Group differences in behavioral GDTs, and CAEP amplitude and latency were evaluated using multiple mixed effects models. Bivariate and multivariate canonical correlation analyses were used to evaluate the relationship between the CAEP amplitude and latency, behavioral GDTs, and speech perception.

**Results:** A significant effect of participant group was not observed for across-frequency GDTs, instead older participants (> 50 yrs.) displayed larger GDTs than younger participants. CI recipients displayed increased P1 and N1 latency compared to NH participants and older participants displayed delayed N1 and P2 latency compared to younger adults. Bivariate correlation analysis between behavioral GDTs and speech perception measures were not significant (*p* > 0.01). Across-frequency canonical correlation analysis showed a significant relationship between CAEP reference condition and behavioral measures of speech perception and temporal processing.

**Conclusions:** CI recipients show similar across-frequency temporal GDTs compared to NH participants, however older participants (> 50 yrs.) displayed poorer temporal processing (larger GDTs) compared to younger participants. CI recipients and older participants displayed less efficient neural processing of the acoustic stimulus and slower transmission to the auditory cortex. An effect of gap duration on CAEP amplitude or latency was not observed. Canonical correlation analysis suggests better cortical detection of frequency changes is correlated with better word and sentence understanding in quiet and noise.

## INTRODUCTION

Temporal resolution, the ability to resolve rapid changes within the acoustic signal over time, is critical for speech understanding, music appreciation and sound localization (Moore 2012; Picton 2013; Rosen 1992). A gap detection paradigm is a standard measure of auditory temporal resolution, which uses an adaptive tracking procedure to determine the shortest gap of silence between two acoustic markers that an individual can detect (Fitzgibbons 1983; Fitzgibbons and Gordon-Salant 1987; Fitzgibbons and Wightman 1982; Formby and Forrest 1991; Formby et al. 1998b; Moore 1985; Moore and Glasberg 1988). Historically, the gap of silence is most commonly bound by acoustic markers that are spectrally identical termed within-frequency gap detection (Lister et al. 2002; Lister and Roberts 2005; Moore and Glasberg 1988). The resulting temporal task is to detect discontinuity of the sound within a single channel i.e., neural activity to pre-gap marker offset compared to neural activity to post-gap marker onset. Within-frequency gap detection thresholds (GDTs) for normal hearing (NH) listeners are typically around 5 ms or less under optimal conditions, e.g., sinusoidal stimuli, long marker durations, and supra-threshold presentation levels (Blankenship et al. 2016; Eddins et al. 1992; Heinrich and Schneider 2006; Oxenham 2000; Penner 1977; Phillips et al. 1998; Plomp 1964). By using pure-tones or narrowband noise, the frequency of the pre- and post-gap acoustic markers can be varied so that they are spectrally distinct, resulting in an across-frequency gap detection paradigm. In natural speech, sounds before and after silent gaps are rarely identical in frequency, therefore across-frequency GDTs might be more realistic and representative measure of temporal resolution. However, a limited number of studies have been completed with across-frequency paradigms, and most were completed with NH participants. Cochlear implant (CI) recipients typically display poorer temporal processing skills, which has been associated with poorer speech outcomes post-implantation (Blankenship et al. 2016; Busby and Clark 1999; Muchnik et al. 1994; Sagi et al. 2009). Therefore, due to the importance of temporal resolution and the similarities between across-frequency gap detection and natural speech, it is reasonable to assume that deficits in across-frequency temporal processing result in poorer speech outcomes.

Across-frequency gap detection paradigms require the listener to detect gaps between acoustic markers that differ in frequency content, which can either be spectrally distinct or contain some frequency overlap (Phillips et al. 1997). In this case, the pre- and post-gap markers activate different locations on the cochlear partition, resulting in different populations of auditory nerve fibers being activated. Thus, the temporal task is no longer a simple onset and offset task but a timing comparison between different neural channels (Hanekom and Shannon 1998). When the markers have no spectral overlap, the relative timing computation must rely on central comparisons of peripheral inputs (Phillips and Hall 2000). Similar to within-frequency conditions, there are many factors that can affect across-frequency GDTs. In general, across-frequency GDTs are consistently higher than within-frequency (Formby et al. 1998a; Formby et al. 1998b; Grose et al. 2001; Phillips and Hall 2000; Phillips et al. 1997) and GDTs increase as the pre-gap marker duration is shortened (Phillips et al. 1998; Phillips et al. 1997). GDTs tend to systematically increase as the frequency separation is increased from a half-octave to an octave (Formby and Forrest 1991; Formby et al. 1996; Neff et al. 1982) and becomes asymptotic for greater frequency separations (Formby et al. 1996). The effect of frequency change direction (i.e., increasing or decreasing) on GDTs is variable. While some studies have shown that increasing the marker frequency from the pre-to post-gap marker resulted in smaller GDTs than decreasing marker frequency (Formby et al. 1996; Heinrich and Schneider 2006; Lister et al. 2002; Lister et al. 2000), others have found no effect of frequency change direction (Formby et al. 1998a). Across-frequency GDTs clearly deteriorate with age (Lister et al. 2002; Lister et al. 2000; Pichora-Fuller et al. 2006; Roberts and Lister 2004). Lister et al. (2002) reported older adults (62-74 yrs.) had significantly poorer across-frequency GDTs compared to middle-aged (40-52 yrs.) and younger adults (18-30 yrs.). In addition, for older adults, across-frequency GDTs increase more sharply for increasing marker frequency disparity compared to younger adults (Lister et al. 2002; Roberts and Lister 2004). Collectively, results suggest that age differences in gap detection are modulated by stimulus complexity, with increased stimulus complexity resulting in more pronounced age differences in across-frequency GDTs (Pichora-Fuller 2003; Pichora-Fuller et al. 2006; Snell 1997).

In NH listeners, across-frequency GDTs are extremely variable. For example, using pure-tone stimuli (4 kHz pre-gap to 2.3 kHz post-gap marker) and randomized marker durations (200 to 300 ms), Grose et al. (2001) reported mean behavioral across-frequency GDTs > 50 ms in a group of young NH listeners (n = 8). Heinrich et al. (2004) also measured across-frequency (2kHz pre-gap to 1 kHz post-gap) pure-tone GDTs in 12 NH young adults using stimuli with a fixed duration of 40 ms but reported much smaller GDTs (Mean = 10.9 ms, SD = 6.8). Lister and colleagues (2007, 2011) reported across-frequency GDTs using narrowband noise (2 kHz pre-gap to 1 kHz post-gap) that were significantly smaller in young adults (Mean = 29.2 ms, Range = 9 to 59 ms) compared to older adults (Mean = 56.0 ms, Range = 21 to 147 ms).

With regard to CI recipients, a limited number of studies have examined across-frequency GDTs, all of which used direct electrical stimulation (Hanekom and Shannon 1998; van Wieringen and Wouters 1999). Hanekom and Shannon (1998) systematically examined GDTs as a function of channel separation in three CI recipients. GDTs were consistently the lowest when the pre- and post-gap markers were presented to the same channel with thresholds between 1 to 4 ms (i.e., within-frequency GDT). As the channel separation between the markers increased, the GDTs increased significantly with large variability across participants. For one CI recipient, across-frequency GDTs were between 10 to 20 ms, however, the other two participants had GDTs from 20 to 70 ms and 100 to 200 ms. Similarly, van Wieringen and Wouters (1999) reported that within-channel GDTs were < 5 ms and across-frequency GDTs were significantly elevated with a greater variability (< 50 ms). Results demonstrate that while some CI recipients have across-frequency GDTs comparable to NH individuals, others display elevated GDTs (Hanekom and Shannon 1998; van Wieringen and Wouters 1999). It is important to reiterate that the two aforementioned studies on across-frequency gap detection were conducted with direct electrical stimulation. While electrical GDTs provide a direct measure of temporal processing impairments within the auditory system without the influence from the device factors, they do not reflect the everyday listening situations of CI recipients. Therefore, it is important to evaluate across-frequency temporal processing using acoustic stimuli, which reflects impairments within the auditory system and signal distortion imposed by the speech processing strategy.

Although it is quick and easy to measure behavioral GDTs in most adults, it can be challenging or impossible to obtain reliable thresholds in pediatric patients or difficult-to-test adults due to factors such as memory, cognition, attention, and motivation. In the United States, pediatric CI recipients can be implanted as young as 9 months of age. Furthermore, approximately 30% to 40% of pediatric CI recipients have additional disabilities limiting their ability to provide reliable behavioral responses (Chilosi et al. 2010). Therefore, an objective measure of temporal processing that does not rely on behavioral responses, would be extremely beneficial. Electroencephalographic (EEG) measure, is an safe, non-invasive, objective measure that has historically been used to evaluate responses from the human auditory system (Harris et al. 2012; Martinez et al. 2013; Picton 2011, 2013; Small and Werker 2012). EEG measures are extremely precise in terms of temporal resolution and can capture millisecond-by-millisecond neural activity, providing the ability to monitor and diagnose temporal processing deficits.

In recent years, electrophysiological gap detection paradigms using cortical auditory evoked potentials (CAEPs) have been used to obtain an objective measure of the neural detection of silent gaps within the auditory cortex (Harris et al. 2012; He et al. 2015; Lister et al. 2007; Lister et al. 2011). The CAEP response is characterized by P1, N1, and P2 peaks at approximately 50, 100, and 200 ms, respectively, after stimulus onset or an acoustic change (intensity, frequency, or silent gap) within a stimulus. While most studies have examined within-frequency temporal processing (Atcherson et al. 2009; Harris et al. 2012; He et al. 2012; He et al. 2013; He et al. 2015; Michalewski et al. 2005; Palmer and Musiek 2013, 2014; Pratt et al. 2005; Pratt et al. 2007), a few have examined across-frequency conditions. Lister et al. (2007) examined both within- and across-frequency electrophysiological gap detection in younger adults with normal hearing using narrowband noise stimuli (2 kHz pre-gap to a 1 kHz post-gap marker) with perceptually weighted gap durations for each participant including a standard (1 ms), sub-threshold (behavioral GDT / 2.4), threshold (behavioral GDT), and supra-threshold (behavioral GDT x 2.4) gap conditions. Across-frequency conditions yielded shorter P2 latency, and increased P1, N1, and P2 amplitude compared to within-frequency conditions (2 kHz pre-gap to 2 kHz post-gap marker). As the silent gap between the pre- and post-gap marker became more salient, CAEP peak latency decreased and amplitude increased. In a follow up study using identical within- and across-frequency electrophysiological gap detection paradigm, Lister et al., (2011) reported older adults displayed delayed P2 latency and increased P1 amplitude compared to younger adults for across-frequency conditions. Additionally, older adults exhibited increased frontal activation when processing across-frequency stimuli compared to younger listeners (Lister et al. 2011). Specifically, a temporal-spatial principle component analysis indicated older adults displayed significantly larger frontal activation for across-frequency conditions at 52, 78, 110, 168, 187, and 220 ms. In contrast, younger adults only displayed increased frontal activation for across-frequency conditions at 51 and 167 ms. To our knowledge, there are no comparable across-frequency electrophysiological studies in CI recipients.

A close association between electrophysiological and behavioral GDTs has been reported (< 10 ms) for normal hearing listeners using within-frequency stimuli (Atcherson et al. 2009; He et al. 2012; Palmer and Musiek 2014; Pratt et al. 2005). In CI candidates and recipients with auditory neuropathy, He et al. (2013, 2015) reported a significant negative correlation between open-set word recognition and within-frequency electrophysiological GDTs (ρ = -0.81, *p* <0.01). However, limited studies have been published using across-frequency paradigms. While Lister and colleagues (2007, 2011) examined across-frequency electrophysiological GDTs, they did not include a measure of speech perception nor were the behavioral and electrophysiological GDTs independent of each other. Similarly, in CI recipients, only behavioral measures of across-frequency temporal processing have been reported, which did not include speech perception or electrophysiological GDTs (Hanekom and Shannon 1998; van Wieringen and Wouters 1999). In addition, the CI studies used direct electrical stimulation which prohibits a direct comparison using identical stimuli and procedures to normal hearing participants.

Due to the importance of temporal resolution for accurate speech understanding, the present study was conducted to examine a behavioral and electrophysiological measure of across-frequency temporal processing and speech perception in CI recipients and NH controls. The purpose of the study was to: (1) evaluate temporal processing using the CAEP response to across-frequency pure-tone stimuli that contained silent gaps; (2) evaluate the relationship between across-frequency temporal processing and speech perception; and (3) explore the relationship among demographic characteristics, speech perception, behavioral GDTs, and CAEP response amplitude and latency. This study is unique because it is the first to evaluate behavioral and electrophysiological across-frequency temporal processing and speech perception in CI recipients. In addition, acoustic stimuli instead of electrical stimulation will be used, which will allow a direct comparison between NH and CI recipients.

## MATERIALS AND METHODS

### Participants

Adult CI recipients (n = 11, Mean = 50.4 yrs., Range = 25.2 to 68.3) and age- and gender-matched NH controls (n = 11, Mean = 49.0 yrs., Range = 24.7 to 68.5) were enrolled in the study. Seven CI recipients were unilaterally implanted (tested only in the CI ear) and four were bilaterally implanted (testing completed in each ear separately). NH individuals were tested in the same ear as their CI age- and gender-match, therefore four completed testing in both ears separately and seven completed testing in only one ear. Altogether, a total of 15 NH and 15 CI ears were included in the study. All CI recipients were implant with a Cochlear Americas Implant System (Cochlear Americas, New South Wales, Australia) and were post-lingually deafened, defined as the onset of bilateral severe-to-profound hearing loss after 3 years of age. CI recipients had at least one year of experience (Range = 1.1 to 10.6 yrs.) to ensure optimization of speech processor settings which typically occurs during the first year post-implantation (Holden et al. 2013). CI recipients reported wearing their speech processor during all waking hours. Differences in the age at test between CI recipients and their NH control ranged from 0 to 4.3 years with a mean age difference of 1.6 years. CI recipients’ device and demographic information is displayed in Table 1. All participants were righted-handed, confirmed with the Edinburgh Handedness Inventory (Oldfield 1971), native speakers of American English, and did not report a history of brain damage, neurologic or psychiatric disorders. The research study was approved by the Institutional Review Board of the University of Cincinnati. Informed consent was obtained from all participants and they were paid for participation.

**Table 1.**
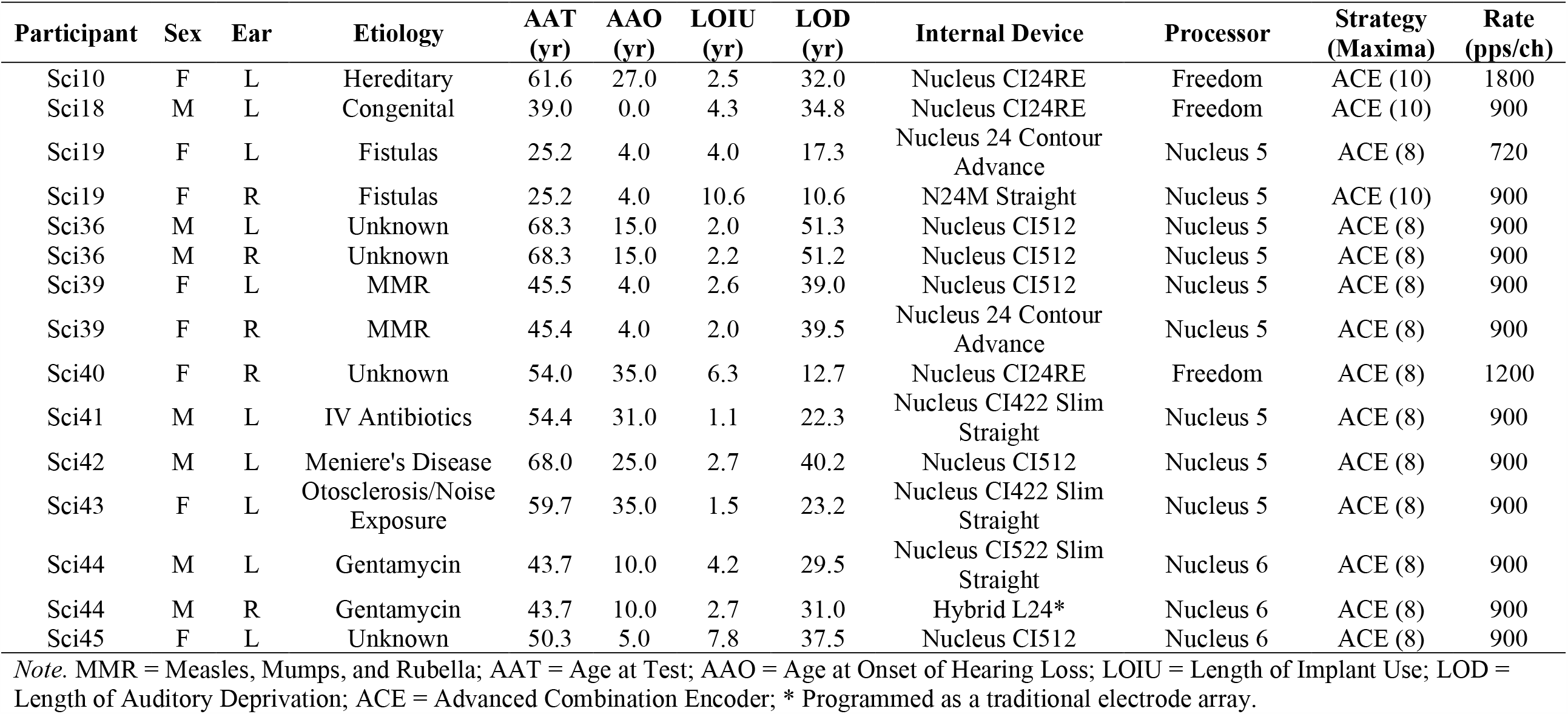
Cochlear Implant Recipient Demographic and Device Information.

## Stimuli

The stimuli for across-frequency behavioral gap detection and electroencephalographic (EEG) recording consisted of a 2 kHz pre-gap pure-tone transitioning to a 1 kHz post-gap pure-tone. The stimuli were created with Audacity software (version 1.2.5; opensource, http://audacity.sourceforge.ent) using a sampling rate of 44.1 Hz. Several studies have demonstrated that pure-tone GDTs are less affected by stimulus complexity (Heinrich et al. 2014; Heinrich and Schneider 2006) resulting in a more accurate measurement of temporal resolution (Moore and Glasberg 1988), therefore pure-tones were used in the study instead of the more common narrow or broadband noise. For behavioral GDT tasks, the pre- and post-gap markers had a variable duration of 250 to 350 ms to prevent participants from using the total duration of the stimulus to aid in behavioral gap detection (Formby and Forrest 1991; Lister et al. 2007). The pre-gap and post-gap markers included a 10 ms rise- and fall-time, respectively. The silent gap durations ranged from 0 to 120 ms (0 ms indicates no gap, 2-20 ms in 2 ms increments, 20-120 in 5 ms increments). The stimuli included a 1-ms rise/fall time around the silent gap to minimize spectral energy spread and were similar to those used in previous studies (Lister et al. 2007; Lister and Roberts 2005; Phillips et al. 1997). The stimuli for EEG testing were identical to those used in behavioral GDT tests, except a wider range of gap durations was included (Range = 1 ms to 360 ms) and the pre- and post-gap marker duration was fixed at 300 ms. The fixed marker duration allowed averaging of the time-locked neural responses across stimulus presentations. Additionally, the longer marker duration of 300 ms enables the separation of the neural responses to the onset of the pre- and post-gap markers.

## General Study Procedures

All testing was completed in the Human Auditory Evoked Potential Lab at the University of Cincinnati within a double-walled sound treated booth (Industrial Acoustics Company, North Aurora, Illinois; (ANSI 1999). The test protocol took between three to six hours to complete, depending on if one ear or both ears were tested. CI recipients completed all testing with their speech processor turned on and adjusted to their everyday settings (volume, sensitivity, and program) which were held constant throughout the test session(s). NH and unilaterally implanted CI recipients completed testing with an E-A-R disposable foam ear plug in the non-test ear to prevent participation. For bilaterally implanted CI recipients, each ear was tested individually with the contralateral speech processor removed. The presentation level for behavioral and CAEP measures was based on individual’s Most Comfortable Level (MCL, 7 on a 0-10 loudness scale; Hoppe et al., 2001) which facilitates comparison of test results between NH and CI participants in current and other studies (Bierer et al. 2015; Dorman et al. 2014; Han et al. 2005). For all participants, behavioral measures were completed first followed by EEG measures. The presentation order of behavioral measures including the lists used within each test, and the order of the CAEP conditions was randomized to minimize order effects.

### Hearing Threshold Assessment

All NH participants completed otoscopy to ensure the ear canal was patent and the tympanic membrane appeared healthy. Next tympanometry was completed with a GSI TympStar Version 2 (Grason Stradler, Eden Prairie, MN) and a traditional 226 Hz probe tone. Audiometric thresholds were measured from 0.25 to 8 kHz using a GSI-61 or AudioStar Pro (Grason Stradler, Eden Prairie, MN) using ER-3A (Etymotic Research, Elk Grove Village, IL) insert earphones. Thresholds were measured using pulsed pure-tones and the Hughson-Westlake procedure (Carhart and Jerger 1959) with a 5 dB step size. Consistent with eligibility criteria, all NH participants displayed normal middle ear status (Compliance = 0.3 to 1.7 ml, Gradient = 50 to 110 daPa, Tympanometric Peak Pressure = -150 to +100 daPa; Jerger, 1970; Margolis & Hunter, 1999) and pure-tone air conduction thresholds that were ≤ 25 dB HL. CI recipients also completed audiometric testing using frequency-modulated tones presented through sound-field speakers to ensure a 25 to 30 dB sensation level (SL) for behavioral and EEG stimuli presented at individual MCL.

### Speech Perception Assessment

NH and CI speech perception performance was measured using the Minimum Speech Test Battery for Adult Cochlear Implant Users. This test battery is comprised of several measures that assess open-set word and sentence recognition in quiet and noise including the Consonant-Nucleus-Consonant Word Test (CNC), Arizona Biomedical Sentence Test (AzBio), and Bamford-Kowal-Bench Speech-in-Noise Test (BKB-SIN; Etymotic, 2005; Peterson & Lehiste, 1962; Spahr et al., 2012). For NH participants, stimuli were presented monaurally through an insert earphone and for CI recipients, stimuli were presented through a loudspeaker at 0 degrees azimuth 1 meter from the CI recipient. Participants were instructed to listen to the speech stimuli and repeat verbatim what they heard, to guess if they were unsure and they did not receive feedback back on their responses. See companion manuscript for additional details on the speech perception measures (Blankenship et al.).

### Temporal Processing Assessment

A psychoacoustic behavioral gap detection task was used to evaluate across-frequency temporal processing abilities. APEX software was used to present stimuli (Francart et al. 2008) with an adaptive two-alternative forced-choice paradigm with an up-down staircase procedure. The participant was presented with two stimuli on each trial, the target stimuli which contained a silent gap and the reference stimuli that did not contain a gap. The order of the target and reference stimuli were randomized for each trial and the duration between trials was set at 0.5 seconds. Participants were instructed to ignore the change in frequency and to select the stimuli that contained the silent gap. Visual feedback was provided for each incorrect and correct response. Testing continued until five reversals were completed and the GDT threshold (ms) was recorded as the mean of the last three reversals.

### Electrophysiological Recording

EEG stimuli were presented and recorded using Neuroscan STIM^2^ software and Neuroscan™ recording system (SCAN software version 4.3, Compumedics Neuroscan, Inc., Charlotte, NC) coupled with a NuAmps digital amplifier. A Neuroscan Quik-Cap (Compumedics Neuroscan, Inc., Charlotte, NC) that contains 40 sintered Ag/AgCl electrodes placed according to the extended 10-20 International system was used to record continuous EEG activity at a sampling rate of 1000 Hz. The recording reference was the earlobe contralateral to the test ear, which has been shown to reduce stimulus artifact for some CI recipients (Liang et al. 2017; McNeill et al. 2007). Electrooculography was recorded using two electrodes placed at the outer canthus of each eye (horizontal) and two electrodes 1 cm above and below the left eye (vertical). Additionally, for CI recipients, the electrodes adjacent to the transmission coil were not used during recording. Impedances were balanced across electrodes and were kept below 5 kΩ. EEG recordings were collected for a total of four gap duration conditions including: (1) threshold (individual behavioral GDT), (2) supra-threshold (behavioral GDT x 3), (3) sub-threshold (behavioral GDT/3), and (4) reference (no gap). Calculated gap durations were rounded up to the nearest integer. Triggers were fixed at the onset of the pre-gap marker and the inter-stimulus interval was set at 0.9 seconds. A minimum of 200 and 400 stimulus trials for each condition were recorded from NH and CI participants, respectively. The stimuli were presented at MCL (7 on a 0-10 loudness scale, Hoppe et al., 2001) through a sound-field speaker to both NH and CI participants. The speaker was 1 meter from test ear at 90 and -90 degrees azimuth corresponding to the right and left ear, respectively. Participants were asked to relax during EEG testing, ignore the stimuli and were instructed to read a self-chosen book or magazine to stay alert. EEG testing took approximately 2 hours per ear to complete and breaks were offered as necessary.

## Data Analysis

### Behavioral Data

The CNC word and AzBio sentence recognition test were scored based on percent correct (CNC-Word, CNC-Phoneme, AzBio-Quiet, and AzBio-Noise). For the BKB-SIN, the SNR-50 was calculated which determines the SNR necessary for the individual to understand 50% of the target words in the sentence. The across-frequency GDT was calculated as the average gap duration (ms) on the last three reversals of the behavioral gap detection task.

### Electrophysiological Data

EEG data were initially processed with Neuroscan software version 4.3, which included a digital band-pass filter (.1 to 30 Hz at 6 dB/octave roll-off), epoched from -100 to 200 ms beyond the offset of the post-gap marker, and baseline corrected using the 100 ms pre-stimulus window. The total epoch duration varied across participants due to the individualized gap durations that were used for the sub-threshold to supra-threshold conditions. Next EEG data was imported into EEGLAB 13.6.5b, an online open source toolbox (http://sccn.ucsd.edu/eeglab) run under Matlab R2017b (The Mathworks, Natick, MA). Unused electrodes surrounding the speech processor coil or those with bad impedances were removed and data were visually inspected to remove epochs that contained non-stereotype artifacts. Independent Component Analysis (ICA) was run to separate the data into neural and artifactual sources. Artifacts arising from ocular movement, electrode or cochlear implant artifact were removed through visual inspection of component properties including the waveform, 2-D voltage map, and the spectrum (Delorme and Makeig 2004; Delorme et al. 2007). Next deleted channels were interpolated, re-referenced to the average reference (Delorme and Makeig 2004; Hagemann et al. 2001), baseline corrected (−100 to 0 ms), and band-pass filtered (.1 to 30 Hz). The CAEP response was most prominent at the central/midline electrodes (Fz, FC3, FCz, FC4, Cz), verified through visual inspection of the scalp topography. Data were averaged across the aforementioned five electrodes to form a single averaged EEG waveform to improve peak identification (Harris et al. 2012; Michalewski et al. 2005). A total of four average waveforms were derived for each participant, one for each gap duration condition (reference, sub-threshold, threshold, and supra-threshold).

CAEP peak components (P1, N1, and P2) were identified for both the pre- and post-gap markers for each averaged waveform. For the pre-gap CAEP, the maximum positive peak between 25 and 75 ms was identified as P1, the maximum negative peak between 75 and 150 ms was identified as N1 and the following positive peak occurring between 150 and 220 ms that was morphologically appropriate was identified as P2 (Harris et al. 2012; Martin 2007). For post-gap CAEP waveforms, peak latencies may vary based on hearing status (NH vs CI) and the saliency of the silent gap (sub-threshold conditions might have increased latency compared to supra-threshold). Therefore, individual latencies for the pre-gap CAEP were used as a guide when identifying post-gap CAEP peak components. CAEP waveforms were visually inspected by two individuals (Blankenship and Zhang) on which they were both in agreement for all CAEP peak components. P1, N1, P2 amplitude was measured as the change in voltage from the baseline to maximum positive or negative peak and the N1-P2 was calculated as the change in voltage from the N1 trough to the P2 peak. For both the pre- and post-gap marker, an average waveform was computed for each gap duration (reference, sub-threshold, threshold, and supra-threshold).

### Statistical Analysis

Outcome variables were evaluated in terms of descriptive statistics and boxplots to evaluate the distribution and identify outliers i.e., values > 1.5 times the interquartile range. A mixed effect model was used to examine differences in across-frequency behavioral GDTs, with participant group (NH vs CI) and test ear (left and right) included as fixed effects and age at test as a covariate. Participant ID was included as a random effect to account for NH and CI participants that were tested in both ears separately. CAEP analyses included mixed effect models that were conducted in a similar fashion, except for an additional fixed effect of condition (reference, sub-threshold, threshold, and supra-threshold). Degrees of freedom were estimated with the Satterthwaite approximation method and the Holm’s step-down adjustment method was implemented to account for multiple comparisons. Several outliers were identified in the data therefore, models were rerun with the outliers removed. If changes in model significance occurred, both analyses were discussed in the results section. While mixed effect models were conducted to examine the effect of group, test ear, and age at test on speech perception scores (CNC, AzBio, BKB-SIN). Results of these analyses are reported in the companion manuscript (Blankenship et al., 2020). Speech perception scores are only included in this manuscript for the purpose of examining the correlation between speech perception and across-frequency temporal processing.

Correlation analysis was conducted using both multivariate canonical and spearman rank correlations. Canonical correlations evaluated the relationship among pairs of variables including CAEP (P1 and N1-P2 amplitude and P1, N1, P2 latency), behavioral (CNC-Word, CNC-Phoneme, AzBio-Quiet, AzBio-Noise, SNR-50, Across-GDT), and demographic variables (age at test, age at onset of hearing loss, length of implant use, length of auditory deprivation). CAEP variables were measured for four different conditions (reference, sub-threshold, threshold, and supra-threshold). This resulted in a total of nine canonical correlation analyses: Behavioral and Demographic (CI data only), Demographic and CAEP (CI data only; reference, sub-threshold, threshold, supra-threshold), Behavioral and CAEP (NH and CI data; reference, sub-threshold, threshold, supra-threshold). Bivariate scatter plots were examined to determine whether the assumptions of linearity, multivariate normality, and homoscedasticity were met. Lastly, one-tailed spearman rank correlation analysis was conducted to evaluate the pairwise relationship between across-frequency behavioral GDTs and speech perception (CNC-Word, CNC-Phoneme, AzBio-Quiet, AzBio-Noise, and SNR-50). All data was analyzed using SPSS statistical software (IBM Corp. Released 2019. IBM SPSS Statistics for Windows, Version 26.0. Armonk, NY: IBM Corp.).

## RESULTS

### Across-Frequency Behavioral Results

Consistent with inclusion criteria, all NH individuals displayed thresholds that were ≤ 25 dB HL from 0.25 to 8 kHz. Mean NH audiometric thresholds are as follows: 0.25 kHz = 12 dB; 0.5 kHz = 13 dB; 1 kHz = 13 dB; 2 kHz = 13 dB; 4 kHz = 15 dB; 8 kHz = 12 dB. In CI recipients, audiometric thresholds ranged from 15 dB HL to 45 dB HL from 0.25 to 6 kHz with the following mean audiometric thresholds (0.25 kHz = 28 dB; 0.5 kHz = 29 dB; 1 kHz = 28 dB; 2 kHz = 25 dB; 4 kHz = 33 dB; 6 kHz = 38 dB). The purpose of obtaining audiometric thresholds in both the NH and CI group was to ensure a 25 to 30 dB SL recommended for optimal gap detection (Fitzgibbons and Gordon-Salant 1987). The NH group had a mean presentation of 48 dB SL (Range = 43 to 55 dB SL) and the CI group had a mean presentation level of 37 dB SL (Range = 25 to 45 dB SL).

Across-frequency GDTs were similar between NH (Mean = 58.8 ms; SD = 38.3; Range = 14.6 to 120.0) and CI recipients (Mean = 82.4 ms; SD = 30.7; Range = 25.0 to 120.0). However, NH participants showed a non-significant trend of lower GDTs. In the NH group, seven ears had GDT < 35 ms (Mean = 25.6 ms, Range = 14.6 to 31.7) with 8 ears displaying GDTs > 45 ms (Mean = 87.7 ms, Range = 46.4 to 120). However, in the CI group, only three ears had GDTs < 35 ms (Mean = 28.9 ms, Range = 25 to 33.3 ms) and 12 ears displayed GDTs > 75 ms (Mean = 95.8 ms, Range = 76.7 to 120). Mixed effect model results did not show a significant effect of group (F[1,18.3] = 3.6, *p* = 0.072), or test ear (F[1,7.1] = 3.7, *p* = 0.094). However, age at test was significant (F[1,18.5] = 18.3, *p* < 0.001), indicating that older individuals (> 50 yrs.; Mean = 90 ms) had larger GDTs than younger participants (Mean = 48 ms).

### Across-Frequency Electrophysiological Results

ICA is a method commonly used to remove CI artifact from EEG recordings. Figure 1 displays across-frequency CAEP waveforms from one CI (top) and one NH participant (middle) prior to ICA and the bottom panel displays both the NH and CI CAEP waveforms after ICA. For all participants in the study, ICA was able to successfully remove the CI artifact to reveal the CAEP waveform. As shown in the figure, there are two-time frames of interest, one after the onset of the pre-gap marker and the second after the onset of the post-gap marker.

**Figure 1.**
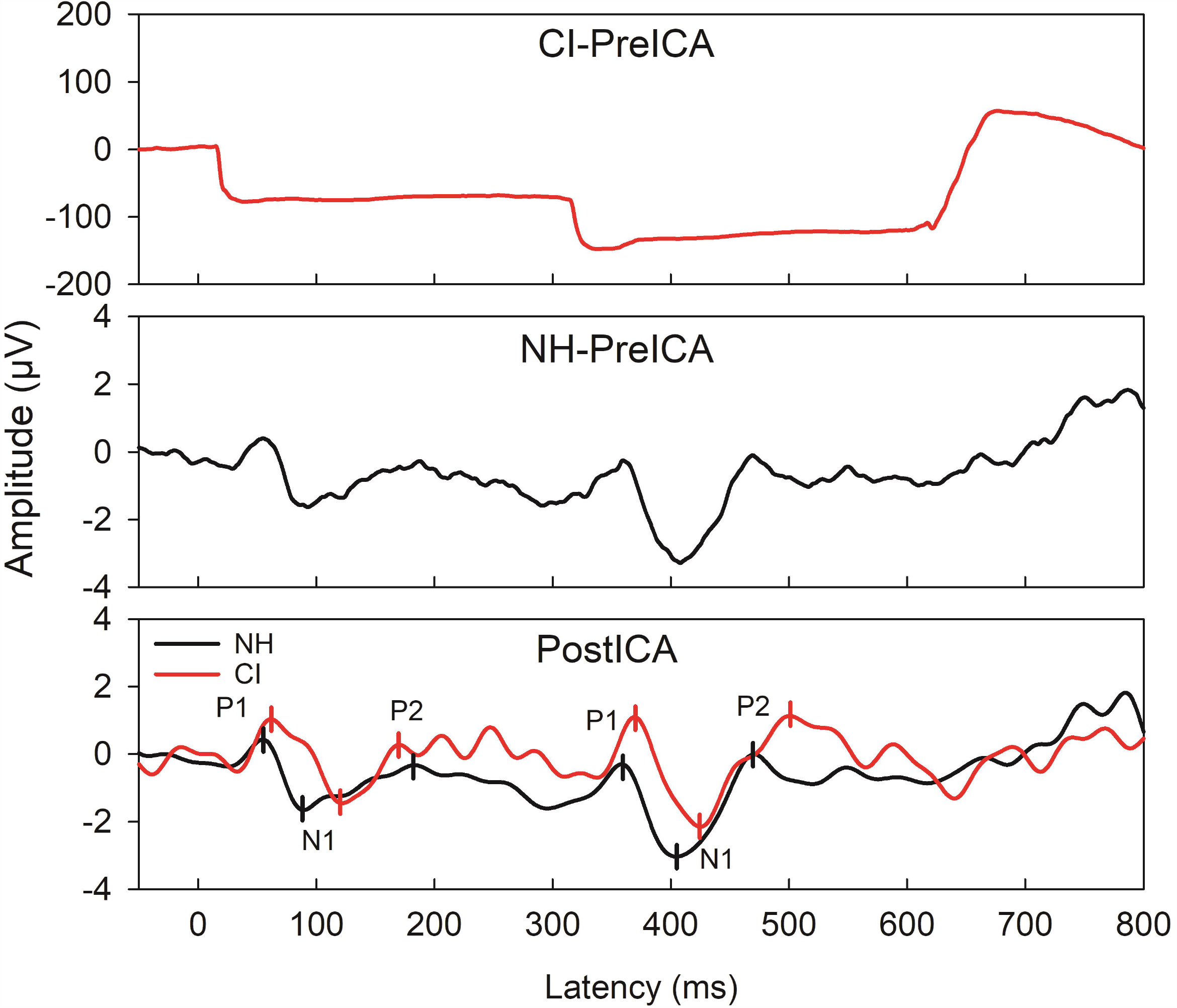
Pre- and post ICA CAEP waveforms from one CI user (Sci45) and their age- and gender-matched NH control for the across-frequency supra-threshold condition. The top two figures display the CAEP before ICA for the CI and NH participant, respectively. The bottom figure displays the NH and CI waveform after ICA, which shows that the pre- and post-gap CAEP are apparent for the CI user. The pre- and post-gap P1-N1-P2 are marked for both the NH and CI participant in the bottom figure. The gap duration was 246 ms for the CI recipient and 81 ms for the NH control.

The pre-gap marker P1-N1-P2 peak components were present for all conditions for both NH and CI participants. Since the pre-gap markers are identical across conditions (i.e., 2 kHz pure-tone), CAEP waveforms were averaged across groups, resulting in one NH and one CI grand average CAEP waveform (Figure 2). Compared to NH participants, CI recipients displayed decreased amplitude (N1 and N1-P2) and increased latency (P1, N1, P2).

**Figure 2.**
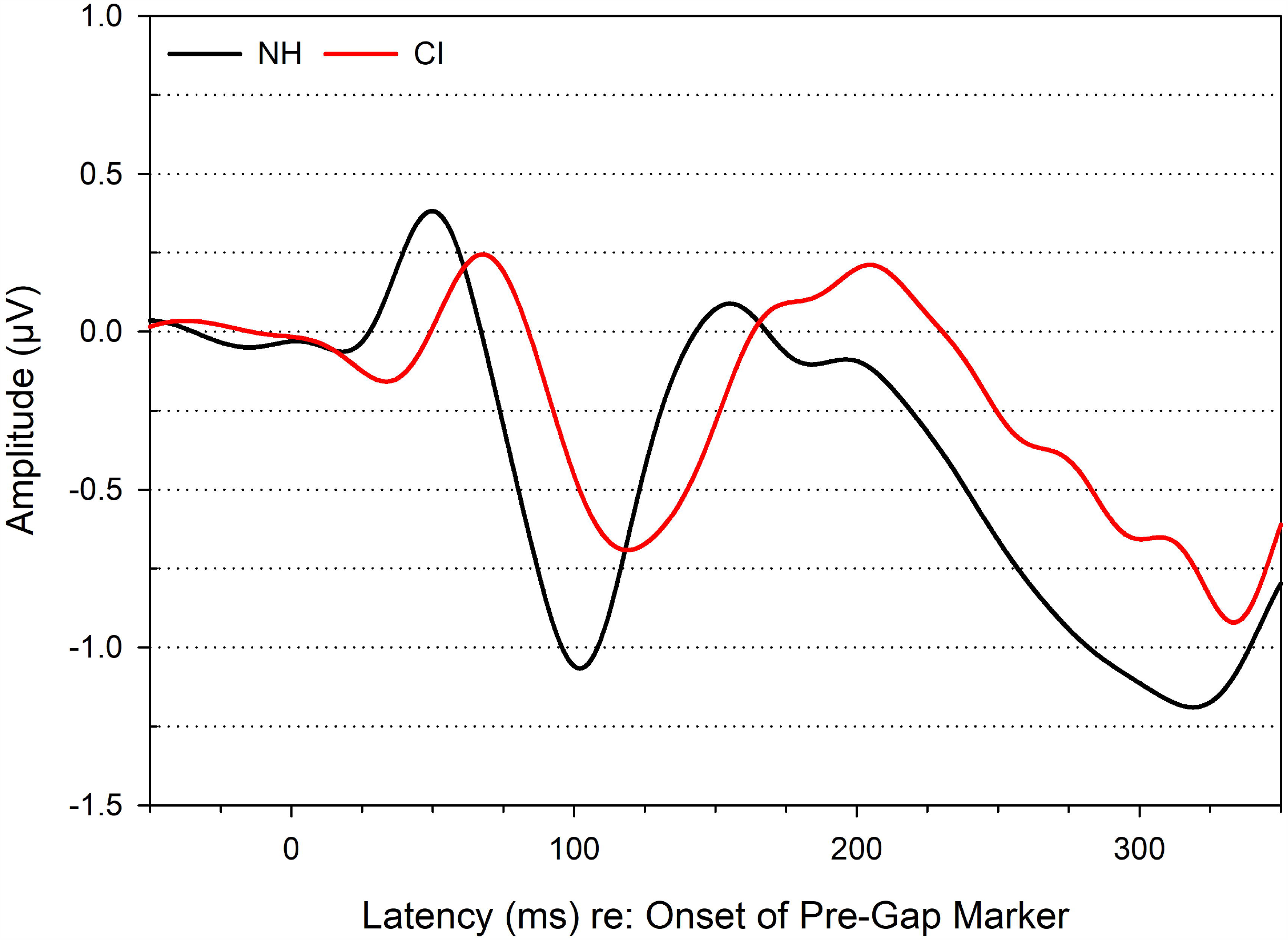
NH and CI group mean pre-gap CAEP waveforms for all across-frequency conditions (reference, sub-threshold, threshold, supra-threshold) averaged together.

Post-gap marker CAEP waveforms are displayed in Figure 3 for each group and condition. Across-frequency CAEPs mean and standard deviation amplitude and latency values along with the number of present CAEPs for each condition are displayed in Table 2. Multiple mixed effect models were used to evaluate the effect of group, condition (reference, sub-threshold, threshold, supra-threshold), test ear, and age at test on CAEP amplitude and latency values (see Table 3). For P1 amplitude, a significant effect of condition was observed (*p* = 0.043). However, after the Holm’s step-down method to adjust for multiple comparisons, a significant difference was not found between any gap duration conditions. No other significant effect of group, condition, test ear, or age at test were observed (*p* > 0.05) for N1, P2, or N1-P2 amplitude.

**Table 2.**
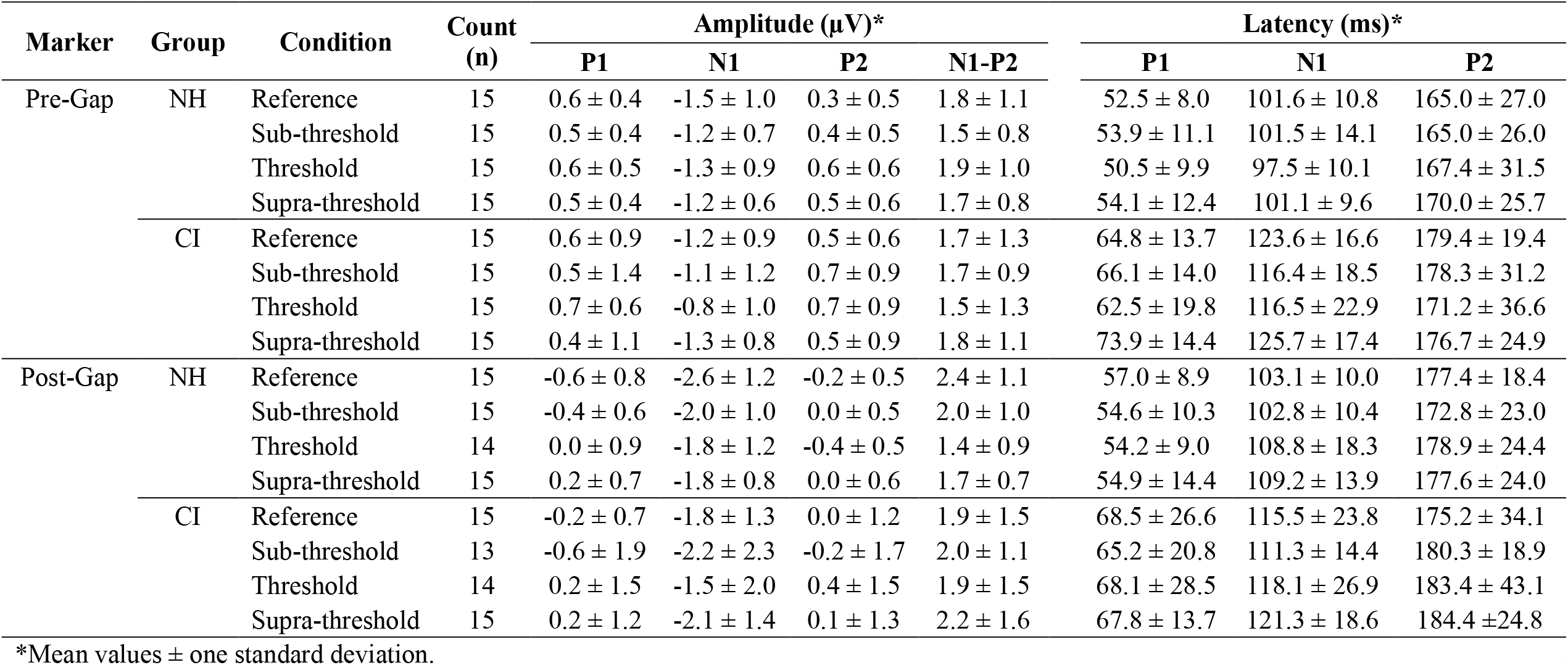
Across-Frequency NH and CI Group Mean CAEP Amplitude and Latency Values

**Table 3.** Across-Frequency Post-Gap CAEP Mixed Effect Model Analysis (*p-*values, F-test, degrees of freedom displayed).

|  | Group | Condition | Test Ear | Age at Test |
| --- | --- | --- | --- | --- |
| P1 Amplitude | 0.690 | ***0.043*** | 0.617 | 0.479 |
| F (DF = 11 to 89) | 0.166 | 2.829 | 0.252 | 0.538 |
| N1 Amplitude | 0.792 | 0.288 | 0.240 | 0.485 |
| F (DF = 17 to 109) | 0.072 | 1.274 | 1.396 | 0.508 |
| P2 Amplitude | 0.279 | 0.935 | 0.156 | 0.475 |
| F (DF = 8 to 87) | 1.297 | 0.141 | 2.050 | 0.560 |
| N1-P2 Amplitude | 0.961 | 0.163 | 0.248 | 0.754 |
| F (DF = 18 to 105) | 0.003 | 1.745 | 1.350 | 0.101 |
| P1 Latency | ***0.017*** | 0.736 | 0.196 | 0.608 |
| F (DF = 15 to 104) | 6.819 | 0.424 | 1.691 | 0.275 |
| N1 Latency | ***0.017*** | 0.139 | ***0.012*** | ***0.010*** |
| F (DF = 14 to 95) | 7.297 | 1.878 | 6.502 | 9.560 |
| P2 Latency | 0.710 | 0.740 | 0.438 | ***0.003*** |
| F (DF = 15 to 108) | 0.143 | 0.418 | 0.606 | 12.171 |

*Note.* Bold italics indicate significant *p*-values.

**Figure 3.**
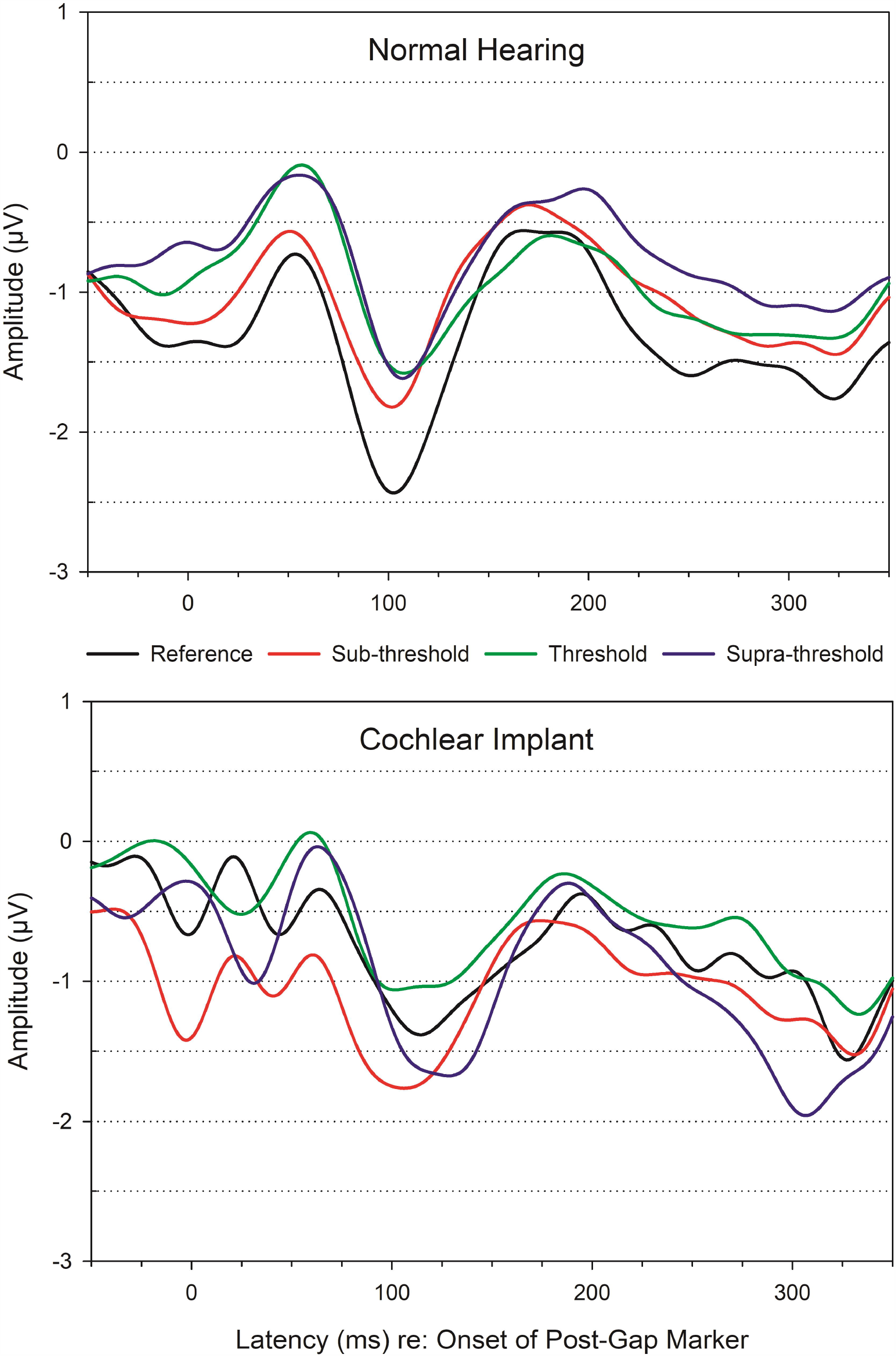
Mean CAEP waveforms for the across-frequency post-gap marker displayed for each gap duration condition in NH (top) and CI groups (bottom).

For CAEP latency values, a significant effect of group was found for P1 (F [1,18.4] = 6.8, *p* = 0.017) where CI recipients displayed slightly longer latencies (Mean = 65.4 ms) compared to NH participants (Mean = 55.9 ms). A significant effect of group (*p* = 0.017), test ear (*p* = 0.012), and age at test (*p* = 0.010) was found for N1 latency. CI recipients (Mean = 117.1 ms) displayed increased N1 latencies compared to NH participants (Mean = 107.8 ms), individuals ≥years of age displayed delayed N1 latencies (younger adults = 95.3 ms, older adults = 99.2 ms), and right ear latencies were increased (Mean = 116.7 ms) compared to left ear responses (Mean = 108.2 ms). Age at test also reached significance for P2 latency (*p* = 0.003) with younger adults (≤ 50 yrs.) displaying a mean P2 latency of 151.1 ms and older adults (> 50 yrs.) displaying a mean P2 latency of 171.4 ms.

Descriptive statistics identified several outliers for across-frequency CAEP amplitude and latency measurements, therefore mixed effect models were rerun with outliers excluded to evaluate their influence on the main effects. With outliers excluded, for P1 amplitude there was no longer a significant effect of condition, instead there was an effect of test ear (F [1,96.3] = 4.4, *p* = 0.039) and age at test (F [1,10.8] = 5.5, *p* = 0.04). The left ear displayed significantly smaller P1 amplitude (Mean = -0.36 µV) than the right ear (Mean = -0.05 µV) and younger adults had significantly smaller P1 amplitude (Mean = -0.36 µV) than older adults (Mean = -0.13 µV). No other changes occurred in mixed model significance values for CAEP amplitude and latency with outliers excluded.

### Across-Frequency Canonical Correlation Analysis

Relationships among CAEP (peak amplitude and latency), Behavioral (CNC-Word, CNC-Phoneme, AzBio-Quiet, AzBio-Noise, SNR-50, and Across-GDT), and Demographic variables (Age at Test, Age at Onset of HL, Length of Implant Use, and Length of Auditory Deprivation) were assessed using multiple multivariate canonical correlations. No significant relationships were found between Demographic and CAEP variables or Demographic and Behavioral variables. For correlation analysis between Behavioral and CAEP variables, only the reference CAEP condition reached significance.

The Behavioral and CAEP reference condition canonical correlation analysis supported a one-dimensional relationship (Wilks’ Lambda = 0.109, F [30, 74] = 2.205, *p* = 0.019). The first canonical correlation coefficient was 0.790 (62% overlapping variance) with an eigen value of 1.658. CAEP threshold coefficients and cross-loadings for the first canonical correlation are displayed in Table 4. Standardized canonical correlation coefficients, which represent each individual item weight to the overall variate, show that CNC-Phoneme, AzBio-Quiet and SNR-50 contribute the most with smaller contributions from CNC-Word, AzBio-Noise, and Across-GDT. For the CAEP variate, N1 latency contributes the most followed by almost equivalent contributions from P2 and P1 latency, with smaller contributions from N1-P2 and P1 amplitude. Cross-loadings show that CNC-Phoneme had the highest correlation value of 0.44 followed by CNC-Word at 0.38 resulting in 19% and 14% of the variance in CNC scores that can be explained by the first CAEP variate. All other speech perception measures (AzBio-Quiet, AzBio-Noise, and SNR-50) and Across-GDT displayed much lower correlations (≤ 0.10) resulting in < 1 percent of the variance in individual measures explained by the first CAEP variate. CAEP amplitude and latency variables displayed similar absolute correlation values ranging from 0.19 to 0.31. The percent of variance in each of the CAEP variables explained by the first behavioral variate is as follows: P1 amplitude (5%), N1-P2 amplitude (8%), P1 Latency (5%), N1 Latency (4%), P2 latency (10%). Overall results indicate a significant relationship between better word understanding in quiet and CAEP amplitude and latency. Specifically, better CNC-Word and CNC-Phoneme scores are related to decreased P1 amplitude, increased N1-P2 amplitude, and increased P1 and P2 latency with decreased N1 latency.

**Table 4.** Across-Frequency Behavioral and Post-Gap Reference CAEP Canonical Correlation Analysis

| Item Content | First Canonical Correlation |  |
| --- | --- | --- |
|  | Cross-Loading | Coefficient |
| **Behavioral** |  |  |
| CNC-Word | 0.38 | 0.79 |
| CNC-Phoneme | 0.44 | 1.44 |
| AzBio-Quiet | 0.10 | -1.37 |
| AzBio-Noise | 0.08 | 0.58 |
| SNR-50 | -0.06 | 1.07 |
| Across-GDT | 0.04 | 0.27 |
| **CAEP** |  |  |
| P1 Amplitude | -0.22 | -0.17 |
| N1-P2 Amplitude | 0.28 | 0.47 |
| P1 Latency | 0.19 | 0.76 |
| N1 Latency | -0.31 | -1.00 |
| P2 Latency | 0.22 | 0.78 |

*Note.* SNR-50 = Signal-to-Noise Ratio required for 50% correct; Across-GDT = Across-Frequency Gap Detection Threshold.

### Across-Frequency Bivariate Correlation Analysis

Non-parametric spearman rank correlation analysis was used to assess the direct relationship between across-frequency GDT and speech perception for all NH and CI participants grouped together (see Figure 4). Results show negative relationships between CNC word recognition and AzBio sentences and across-frequency GDT and a positive relationship between BKB-SIN SNR-50 and across-frequency GDT. However, after Bonferroni correction to account for multiple comparisons (n = 5; *p* ≤ 0.01), none of the correlations reached significance.

**Figure 4.**
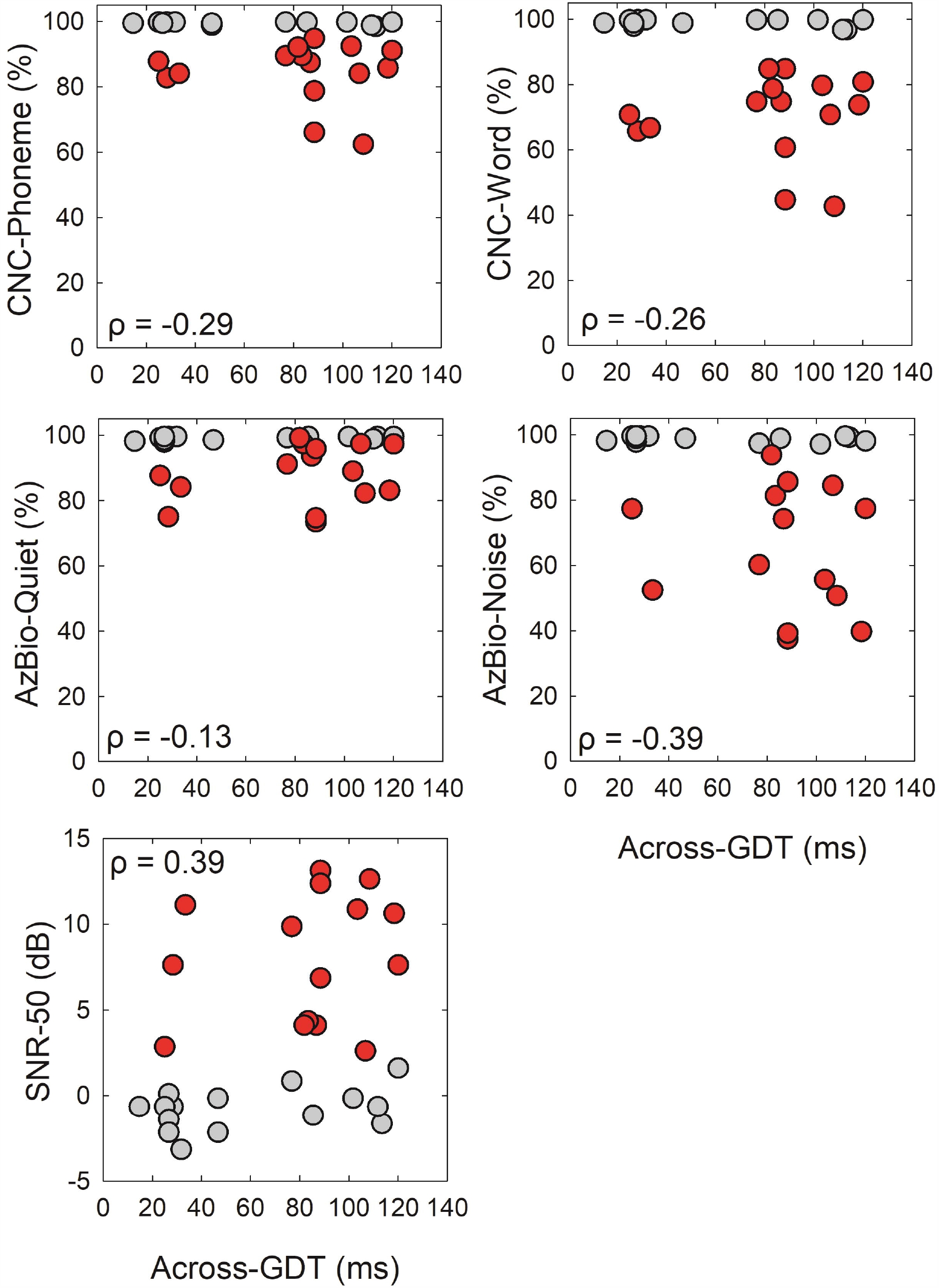
Speech perception scores plotted as a function of across-frequency GDTs. NH and CI data points are displayed in grey and red, respectively. Results of a spearman’s ranked correlation test are displayed in each graph. After a Bonferroni correction, none of the correlations were significant (p > 0.01).

## DISCUSSION

The purpose of the present study was to evaluate across-frequency temporal processing using both behavioral and electrophysiological measures in CI recipients and age- and gender-matched NH controls and examine the correlation with speech perception. NH and CI recipients performed similarly on across-frequency gap detection, but older individuals (> 50 yrs.) had significantly higher GDTs than younger individuals. Mixed effect model analysis showed no significant effect of group (after pairwise comparison correction), condition, test ear, and age at test on CAEP amplitude. For CAEP latency, multiple significant effects of group, test ear, and age at test were found with no significant effect of condition. Canonical correlations showed a significant correlation between across-frequency CAEP reference condition (i.e., no gap) and behavioral speech perception and temporal processing. After a Bonferroni correction, non-parametric spearman rank correlation analysis did not reveal a significant relationship between across-frequency behavioral GDTs and speech perception.

### Across-Frequency Behavioral Gap Detection

NH and CI recipients displayed a non-significant mean difference in across-frequency GDTs (NH = 58.8 ms, CI = 82.4 ms) with both groups displaying wide variability in thresholds. The NH group mean GDTs are consistent with previous studies that have documented wide variability in across-frequency GDTs (Formby et al. 1998a; Formby et al. 1998b; Grose et al. 2001; Heinrich et al. 2004; Lister et al. 2007; Lister et al. 2011; Phillips and Hall 2000; Phillips et al. 1997). Grose et al. (2001) reported across-frequency GDTs > 50 ms for young NH adults. Heinrich et al. (2004) reported smaller across-frequency GDTs (Mean = 10.9 ms) using 2 kHz to 1 kHz pure-tone stimuli and shorter marker durations. Using 2 kHz to 1 kHz pre-to post-gap narrowband noise and a similar paradigm to the current study, Lister et al. (2007, 2011) reported across-frequency GDTs that ranged from 9 to 59 ms (Mean = 29.2 ms, SD = 4.3) in young adults and GDTs that ranged from 21 to 147 ms (Mean = 56.0 ms, SD = 6.1) in older adults. More recently, Alhaidary and Tanniru (2019) reported across-frequency narrowband noise GDTs in NH adults (Mean = 36.0 ms, SD = 14.9) which were comparable to those reported by Lister et al. (2007) but are still significantly lower than the GDTs obtained in NH adults in the current study. The standard deviation in the current study for NH participants was 38 ms, which is much higher than previously reported (Heinrich et al. 2004; Lister et al. 2007; Lister et al. 2011). The discrepancy between across-frequency GDTs obtained in the current study and previous studies could be due to differences across studies regarding participant age, instrumentation, stimulus, and presentation levels.

There are only two across-frequency GDT studies in the literature involving CI recipients, both conducted with direct electrical stimulation. Hanekom and Shannon (1998) measured across-frequency GDTs using 200 µs/phase biphasic pulses with a stimulation rate of 1000 pps in three adult CI recipients (Range = 39 to 55 yrs.) as a function of electrode separation (Range = 1 to 18 electrodes). Across-frequency GDTs systematically increased as the channel separation increased with thresholds varying significantly across participants (Range = 10 to 200 ms). Upon examining across-frequency GDTs using electrode pairs corresponding to the frequencies used in the current study, GDTs were approximately 7, 25, and 50 ms in these 3 participants (Mean = 48.7 ms). van Wieringen and Wouters (1999) reported similar across-frequency GDTs in four post-lingually deafened CI recipients (Mean = 32 yrs., Range = 13 to 46) using 40 to 100 µs/phase biphasic pulses with a stimulation rate of 1250 pps. Half of the participants displayed very low across-frequency GDTs (<10 ms), while the other two had higher GDTs (10 to 30 ms). In the current study, mean across-frequency GDTs in the CI group was 82.4 ms (Range = 25 to 120 ms), which is higher than most individual GDTs reported in previous studies (Hanekom and Shannon 1998; van Wieringen and Wouters 1999). The large differences in across-frequency GDTs is likely due to differences in stimulus presentation mode across studies. The current study used acoustic stimulation while the previous studies used direct electrical stimulation. Acoustic GDTs reflect the temporal processing impairments within the auditory system and signal distortion associated with the speech processing strategy and individual map parameters (stimulation rate, maxima, preprocessing strategies). Therefore, acoustic GDTs are expected to be poorer than those measured with direct electrical stimulation and are more representative of temporal processing abilities in everyday situations.

Lastly, a significant effect of age at test was found for across-frequency GDTs, where older individuals (> 50 yrs., CI and NH combined) had significantly larger GDTs (Mean = 90.3 ms, SD = 30.1) than younger individuals (Mean = 48.1 ms, SD = 29.1). This finding is consistent with studies by Lister and colleagues (2007, 2011), who also reported significantly larger across-frequency GDTs in older individuals with normal hearing or minimal hearing loss. Furthermore, across-frequency GDTs obtained in the current study were significantly larger than GDTs reported by Lister et al. for older (Mean = 56.0 ms) and younger adults (Mean = 29.2 ms). This could be due to the inclusion of CI recipients in the current study. When GDTs are examined as a function of group and age at test, CI recipients displayed mean GDTs that were much higher (Younger = 65 ms, Older = 98 ms) than in the NH group (Younger = 31 ms, Older = 83 ms). While the difference is more pronounced for younger participants, we believe this may have contributed to the poorer overall across-frequency GDTs in the current study compared to Lister and colleagues.

### Across-Frequency Electrophysiological Gap Detection

Across-frequency CAEPs were recorded using pure-tone stimuli (2 kHz pre-gap to 1 kHz post-gap) and four gap duration conditions including: (1) reference (no gap), (2) sub-threshold (behavioral GDT/3), (3) threshold (behavioral GDT), and (4) supra-threshold (behavioral GDT x 3). With regard to post-gap CAEP amplitude, mixed effect model analysis initially showed that P1 amplitude was significantly affected by gap duration (*p* = 0.043). Mean P1 amplitudes were slightly smaller for the no gap and shorter gap duration conditions (reference = -0.3 µV, sub-threshold = -0.5 µV) compared to the longer gap duration conditions (threshold = 0.1 µV, supra-threshold = 0.2 µV). However, after the Holm’s step-down method to adjust for multiple comparisons, a significant effect of gap duration was no longer observed (*p* > 0.05). No other significant effects of group, gap duration condition, test ear, or age at test were observed for post-gap CAEP amplitude. A limited number of studies have examined across-frequency electrophysiological GDTs that can be used for comparison purposes, all of which were conducted with individuals with normal or minimal hearing loss (Lister et al. 2007; Lister et al. 2011). Lister and colleagues (2007, 2011) examined across-frequency temporal processing using narrowband noise stimuli (2 kHz pre-gap to 1 kHz post-gap marker) in a group of young and older adults. Similar to the current study they included three gap durations conditions that were based on the behavioral GDTs including a threshold (behavioral GDT), sub-threshold (behavioral GDT / 2.4), and supra-threshold (behavioral GDT x 2.4) condition. However, instead of using a reference condition (no gap) as in the current study, they used a standard condition which contained a 1 ms gap. Results showed young NH listeners (Mean = 26 yrs., Range = 21 to 40) had a significantly smaller P1 amplitude for the standard condition (.3 µV) than for threshold (1.1 µV) and supra-threshold condition (.7 µV), and the sub-threshold condition P1 amplitude (.4 µV) was significantly smaller than the threshold condition (Lister et al. 2007). Older adults (Mean = 63 yrs., Range = 55 to 74) with normal hearing or minimal hearing loss displayed P1 amplitudes that were significantly larger for the supra-threshold condition (1.4 µV) than for all other gap duration conditions; standard = 0.7 µV, sub-threshold = 0.8 µV, threshold = 1.0 (Lister et al. 2011). When looking at the effect of age on CAEP amplitude, (Lister et al. 2011) reported a significantly larger P1 amplitude in older compared to younger adults (*p* = 0.05). Since P1 attenuation reflects the inhibition of attention to irrelevant stimuli whereas enhancement reflects increased attention to task-relevant stimuli, they proposed that younger listeners are better able to ignore irrelevant stimuli. In contrast to Lister and colleagues, no significant effects of gap duration condition or age at test on CAEP amplitude were found in the current study. Furthermore, mean P1 amplitude across gap duration conditions in the current study ranged from -0.5 to 0.2 µV, which is considerably smaller than the P1 amplitudes reported by Lister et al. for younger (Range = 0.3 to 1.1 µV) and older adults (Range = 0.7 to 1.4 µV).

The absence of significant differences in post-gap CAEP amplitude (Group and Age at Test) observed in the current study might be due to the inclusion of both older adults and CI recipients, populations with known temporal processing impairments. Differences may also be attributed to the availability of neural resources to encode gaps. Animal studies have shown older animals can have up to 50% less cortical neurons available to encode temporal information (Brody 1955; Walton et al. 1998) and CI recipients can have a reduced number of cortical neurons due to auditory deprivation. Therefore, these two factors in conjunction could partially account for the reduced CAEP amplitudes compared to previous studies. Alternatively, the larger amplitudes reported by Lister et al., (2007, 2011) could be due to the use of quarter-octave narrowband noise, exciting a larger area on the basilar membrane which may contribute to the larger overall CAEP amplitude. Lastly, since smaller P1 amplitudes are thought to reflect the inhibition of attention to irrelevant stimuli, individuals in the current study might be better at disregarding irrelevant stimuli.

With regard to CAEP latency, CI recipients displayed significantly increased P1 (NH = 55.9 ms, CI = 65.4 ms) and N1 latency (NH = 107.9 ms, CI = 117.1 ms) compared to NH participants with no group differences found for P2 latency. The P1 latency in NH participants is similar to previous studies with younger (Mean = 54.2 ms) and older adults (Mean = 56.5 ms; Lister et al., 2007; Lister et al., 2011) while the CI group mean P1 latency was considerably longer. The N1 latency in CI participants is similar to values reported by Lister and colleagues in younger (Mean = 114.1 ms) and older (Mean = 115.2 ms) with NH participants displaying slightly shorter N1 latency (Mean = 107.9 ms). Since P1 reflects the cumulative synaptic delay from the peripheral to central auditory system (Eggermont et al. 1997; Steinschneider et al. 1994), increased latency indicates less efficient neural processing of the acoustic stimulus and slower transmission to the auditory cortex. Additionally, the N1 response is thought to primarily reflect stimulus characteristics including timing and intensity (Naatanen and Picton 1987). Therefore, increased N1 latency may be attributed to the complex nature of the stimulus that includes a change in frequency.

Individuals < 50 years of age displayed decreased mean N1 latencies (Mean = 95.3 ms) compared to older individuals (Mean = 99.2 ms). The same effect of age at test was found for P2 latency, with shorter latencies for individuals < 50 yrs. (Mean = 151.1 ms) compared to older individuals (Mean = 171.4 ms). Similarly, Lister and colleagues (2007, 2011) reported longer P2 latencies using across-frequency stimuli for older adults (Mean = 194.7 ms) compared to younger adults (Mean = 171.8 ms). Collectively, these studies indicated that older adults have delayed neural transmission which negatively impacts gap detection ability and younger adults are more efficient at encoding across-frequency stimuli.

A significant effect of test ear was found for N1 latency, where the right ear displayed a slightly increased latency (Mean = 116.6 ms) compared to the left ear (Mean = 108.2 ms). Since CI recipients and their NH controls were tested in the same ear, the delayed right ear latency can’t be attributed to an increased number of CI participants tested in the right compared to the left ear (i.e., CI recipients had longer latencies than NH individuals). Furthermore, there were an equal number of males and females that were tested in the left and right ear. Thus, differences in head size circumference between participants, which impacts volume conduction and the time it takes to reach generation sites (Aoyagi et al. 1990; Shetty and Puttabasappa 2012) is unlikely to cause a delayed right ear N1 latency as well. The mean age at test was similar for the left (Mean = 50.6 yrs.) and right ear (Mean = 47.5 yrs.). However, the length of auditory deprivation for CI recipients tested in the right ear was slightly shorter (28.9 yrs.) compared to the left ear (32.7 yrs.) but the difference is minimal. Thus, significant ear differences were also unlikely to be caused by differences in age at test or auditory deprivation.

Previous studies have reported a left hemisphere advantage for processing temporal information across behavioral (Nicholls and Whelan 1998), electrophysiological (Liégeois- Chauvel et al. 1999; Nicholls et al. 1999), and anatomical and imaging studies (Musiek and Reeves 1990; Penhune et al. 1996; Zaehle et al. 2004). Behavioral GTDs likely reflect temporal processing abilities in the contralateral hemisphere (Efron et al. 1985), therefore a left hemisphere advantage should result in a right ear superiority. In addition, across-frequency temporal processing is thought to contribute to phoneme identification (Elangovan and Stuart 2008; Phillips and Smith 2004; Phillips et al. 1997), and thus better performance in the right ear would be consistent with an expected left hemisphere advantage. In contrast, the current study showed a right ear N1 latency that was increased compared to the left ear which does not reflect a left hemisphere advantage. Due to inconstancies across studies, Carmichael et al. (2008) proposed that while temporal processing abilities may differ between ears, the difference is small at best. Some have suggested that the left hemisphere advantage manifests as improvements in processing “efficiency” (Hill et al. 2004; Stuart 2008; Stuart et al. 2006) or integration times (Ishigami and Phillips 2008) rather than a right ear advantage on traditional gap detection tasks. A potential alternative explanation is that since the across-frequency task is perceptually dominated by the change in frequency not the silent gap, the shorter latency observed in the left ear might support a right hemisphere dominance. This is consistent with previous studies showing a right hemisphere preference for processing a change in frequency (Zatorre et al. 2002).

### Across-Frequency Canonical Correlation Analysis

Multivariate canonical correlation analysis revealed a significant relationship (62% overlapping variance) between the reference CAEP condition and behavioral measures of speech perception and temporal processing. The significant relationship is primarily between better performance on CNC words and decreased P1 amplitude, increased N1-P2 amplitude, and increased P1 and P2 latency with decreased N1 latency. Minimal contributions are from AzBio, SNR-50 and Across-GDT which all explain < 1% of the variance in CAEP amplitude and latency. A few other studies have reported significant relationships between speech perception and CAEPs to gaps in CI recipients and individuals with auditory neuropathy (He et al. 2013; He et al. 2015; Michalewski et al. 2005). However, all of those studies used within-frequency stimuli, and the CAEP values included were gap detection thresholds (ms) and not P1-N1-P1 peak amplitude and latency values. To our knowledge, there are no other published studies that have investigated the relationship between across-frequency CAEP gap detection and behavioral measures of temporal processing and speech perception. However, since the reference CAEP condition did not contain a silent gap but rather a change in frequency (2 kHz to a 1 kHz pure-tone), studies that examine the ability to detect changes in frequency can be used for general comparison purposes. Frequency change detection tasks have been evaluated in CI recipients previously using both behavioral psychoacoustic task (Zhang et al. 2019) and CAEPs (Liang et al. 2018). In adult CI recipients, Zhang et al. (2019) measured frequency change detection thresholds using a behavioral psychoacoustic task. The stimuli were 1 second pure-tones (.25, 1, and 4 kHz) that contained an upward frequency change at the midpoint with step sizes ranging from 0.5% to 200%. Results revealed a strong significant correlation between frequency change detection thresholds and speech perception (CNC, AzBio-Q, AzBio-N; *p* < 0.001) where lower thresholds result in higher speech perception performance. Additionally, high correlation coefficients (R^2^ = 0.71 to 0.74) indicate that a large portion of the variability in speech perception can be attributed to the variability in spectral resolution abilities. Liang et al. (2018) examined the correlation among cortical and behavioral measures of frequency change detection and speech perception in adult CI recipients. Frequency change detection thresholds were measured using a 0.16 kHz pure-tone with an upward frequency change (0.5 to 65%) at stimulus midpoint (500 ms). The EEG stimuli were.16 and 1.2 kHz pure-tones containing upward frequency changes of 0, 5, and 50%. The CAEP evoked by the frequency change showed good agreement with the behavioral frequency change detection thresholds (thresholds > 5% had a present CAEP response). A significant positive correlation was found between the CAEP N1 latency and frequency change detection thresholds, where smaller thresholds resulted in shorter N1 latency (R^2^ = 0.23, *p* < 0.05). Lastly, a significant correlation was reported between CNC words and CAEP N1 latency (R^2^ = 0.36, *p* < 0.05). Collectively results indicate CAEPs evoked by a change in frequency are correlated with speech perception abilities and can be used as an objective indicator of frequency change detection. In this study, the stimulus for the reference condition is similar to the stimulus containing a frequency change at the midpoint. Therefore, the correlation between the CAEP for the reference condition and the speech perception observed in this study may reflect that the cortical detection of frequency changes is critical for speech perception.

### Across-Frequency Bivariate Correlation Analysis

Temporal cues in natural speech occur between sounds of various spectral compositions. Therefore, it is reasonable to assume that across-frequency behavioral GDTs exhibit a relationship with behavioral speech perception. Previous studies have tested this relationship using phonetic boundaries along the voice-onset time continuum. Elangovan and Stuart (2008) showed a significant positive correlation between voice onset time phonetic boundaries and across-frequency GDTs (1 kHz: r = 0.54, *p* = 0.021; 2 kHz: r = 0.77, *p* < 0.001; 4 kHz: r = 0.57, *p* = 0.014; 8 kHz: r = 0.74, *p* < 0.001). In contrast, Mori et al. (2015) did not find a significant correlation between voice onset time boundaries or slope (/ba/ to /da/) and across-frequency GDTs in a group of Japanese listeners. The current study used clinical measures of speech perception in quiet and noise to assess the relationship with across-frequency GDTs. Results did not reveal a correlation between CNC, AzBio, or SNR-50 and across-frequency GDTs (*p* > 0.01). The relationship between clinical speech perception measures and across-frequency GDTs have not been reported previously. Although temporal processing clearly plays a role in speech perception, our results suggest that there is not necessarily a direct relationship between across-frequency temporal processing and clinical measures of speech perception.

### Implications and Limitations

The across-frequency CAEP response elicited by silent gap durations that range from perceptually inaudible (sub-threshold) to audible (supra-threshold) do not show a correlation with behavioral GDTs or speech perception. However, the reference CAEP condition, which does not contain a silent gap but rather a frequency change, shows a significant relationship with speech perception in quiet (CNC) and noise (AzBio and BKB-SIN). Furthermore, behavioral across-frequency GDTs do not show a significant correlation with clinical measures of speech understanding. Due to the lack of correlation with speech perception and the large discrepancy between behavioral GDTs and CAEP responses, there is limited clinical application of across-frequency gap detection as a measure of temporal processing. However, due to the significant correlation between the CAEP reference condition and speech perception in the current and previous studies (Liang et al. 2018; Zhang et al. 2019), CAEPs evoked by frequency changes might be clinically used to assess spectral resolution.

## CONCLUSIONS

NH and CI participants performed similarly on an across-frequency behavioral gap detection task but poorer GDTs were observed for individuals > 50 years of age compared to younger participants. In contrast to previous studies, the effect of gap duration was not observed for CAEP amplitude or latency. Instead CI recipients displayed delayed P1 and N1 latency relative to NH peers and older adults had increased N1 and P2 latency relative to younger adults. Canonical correlation analyses showed a significant relationship between the across-frequency CAEP peak amplitude and latency for the reference condition and behavioral scores. Since the reference condition, does not contain a silent gap, only a change in frequency, results suggest better cortical detection of frequency changes is correlated with better word and sentence understanding in quiet and in noise. No other significant relationships were found between demographic, behavioral, and CAEP variables. Due to the lack of correlation among electrophysiological conditions that contain a silent gap (sub-threshold through supra-threshold), behavioral across-frequency GDTs and speech perception, there is limited clinical application of across-frequency gap detection as a measure of temporal processing.

## Data Availability

Data can be available upon request

## Acknowledgements

The authors thank all participants in this research. This research was supported by the National Institute Health, National Institute on Deafness and Other Communication Disorders Grant R15 DC011004 (F.Z) and the University of Cincinnati University Research Council (C.M.B). Portions of this article were presented at the Association for Research in Otolaryngology 37^th^ Annual MidWinter Meeting in 2014 and the 42^nd^ Annual MidWinter Meeting in 2019. This manuscript represents a portion of the doctoral dissertation of the first author under guidance of the co-authors. The content is solely the responsibility of the authors and does not necessarily represent the official views of the National Institutes of Health or the University of Cincinnati.

## Author Contributions

C.M.B designed and performed the experiments, analyzed the data and wrote the paper. J.M provided statistical analysis. F.Z. designed the experiments and provided interpretive analysis and critical revision to the paper.

## Abbreviations

AzBio: Arizona Biomedical Sentence Test
BKB-SIN: Bamford-Kowal-Bench Speech-in-Noise Test
CAEP: Cortical Auditory Evoked Potential
CI: Cochlear Implant
CNC: Consonant-Nucleus-Consonant Word Test
EEG: Electroencephalographic
GDT: Gap Detection Threshold
ICA: Independent Component Analysis
MCL: Most Comfortable Level
NH: Normal Hearing
SL: Sensation Level
SNR: Signal-to-Noise Ratio

